# Factors Influencing Health Impacts among Smartphone Users in New Normal Situation: A Case Study among University Students in Thailand

**DOI:** 10.1101/2022.04.26.22274312

**Authors:** Wattasit Siriwong, Eric G. Frost, Wachiraporn Wilaiwan

## Abstract

**Background:** Nowadays, smartphone users are increasing across the world. Smartphones have become a necessary thing in people’s life. Using smartphones have both benefits and health effects. Therefore, this study mainly aims to develop an appropriate mobile application to be the tool for health effects and finding factors associated with a health risk from smartphone usage.

**Methods:** This study was a cross-sectional descriptive study. The data was conducted in Thailand. The sample size was 500 university students. The Smartphone U Health mobile application was developed and shown in the Play Store and App Store. The self-administration was used for data collection throughout the Smart U Health. Data were entered and analyzed with licensed SPSS version 22.

**Results:** The participants, including 328 females (65.6%) and 172 males (34.4%) with an average age was 20.3±1.5 years old. The average period of using smartphones was 7.9±2.0 years. The average time spent was 8.6±3.4 hours/day. The primary physical, mental, and social health effects from smartphone usage over three months were eye pain (93.8%), feeling bored (90.4%), the participants had a communication problem with other people (84.0%), respectively. In the multivariate analysis, the significant factors associated with a health risk from smartphone usage were faculty, income, using smartphones in the classroom, experience of resting their eyes before continuing. Moreover, knowledge and practice levels were significantly associated with health risk levels from smartphone usage.

**Conclusion:** Overuse of smartphones had many health effects. The Smart U Health mobile application might be the appropriate tool to assess smartphone users’ health risk levels. In the future, it may consider an intervention study to reduce health risks from smartphone usage.

## Background

The smartphone is a mobile phone with clearly an evolution that combines many gadgets or appliances such as mobile phone, tracker, TV receiver, sound recorder, video camera, video player, music system, e-book reader, newsstand, Internet navigator, and portable office [1]. Nowadays, smartphone users are increasing across the world. Worldwide, the number of smartphone users were three billion in 2018. This number is expected to increase to 3.8 billion in 2021 [2]. In Thailand, the number of smartphone users has been rapidly increasing since 2015 and reached 52.71 million users in 2020 [3]. The largest Internet usage group in Thailand was the youth group ages between 15-24 years [4]. The number of undergraduate students in Thailand was 1,681,149 [5]. Some of those people who were the most significant use of the internet by smartphone were undergraduate students, mostly between the ages of 18-21 years old [6]. Therefore, a study among university students and smartphone users should be a concern.

The smartphone can be used in everyday life through different activities. Smartphones help people easily communicate via text or voice message between two or more people, or any features that help people stay connected are highlight features of smartphones [7]. Moreover, smartphones can be used for entertainment purposes, such as watching movies and listening to music [8]. Social networking was highlighted as a factor associated with smartphone addiction among every age group and young people [9,10]. Other activities with smartphone use are Internet gaming and gambling, which were considered a feature that enables a behavioral addiction to smartphones [11].

However, the increase in smartphone usage has become an important issue in public health. Using a smartphone induced increased upper extremity muscle activity and caused greater upper trapezius (UT) pain [12]. Moreover, overuse of smartphones can cause adverse such as neck problems [13], hand pain [14]. Other effects of smartphones were traffic and pedestrian accidents [15]. Further, smartphone users who have overused smartphones had decreased face-to-face social relationships and social activity [16]. Even decreased sleepiness can occur by blue light in the smartphone [17]. In Thailand, a few studies are focusing on smartphone usage and health risk. A previous study showed that smartphone and tablet usages positively and negatively associated with physical, mental, and social health effects among older adults in Thailand. Many factors were associated with those health effects from smartphone and tablet usages [10,18].

However, there is a benefit of smartphone usages as the World Health Organization defined mobile health (mHealth) as “medical and public health practice supported by mobile devices, such as mobile phones, patient monitoring devices, personal digital assistants, and other wireless devices”[19]. For both physical and mental health care, smartphone software applications, or “apps” have a variety of functions which can be useful for medicine areas [20]. A previous study found that over 165,000 health-focused apps were available for people to download on their own devices [21]. Therefore, using the mobile application as a tool to study is interesting.

There are limits to the study about smartphone usage status and health risk level from smartphone users in other age groups, mainly from the university students in Thailand, consisting of adolescents and youth, the group with the most significant smartphone use users. Therefore, the study of smartphone usage patterns among university students is essential. Moreover, the health risk of smartphone usage among university students should be of concern. This study is to figure out smartphone usage patterns and survey the health effects of smartphone usages among university students in Thailand via a mobile application. Moreover, aim to find factors associated with a health risk from smartphone usage.

## Methods

### Study Site and Population

The study design in this study was a cross-sectional descriptive study. The data collection period was between August to October 2020. The study area was over Thailand. The participants were the university students studying for bachelor’s degree living in Thailand with Having a smartphone and apply it more than six months, male or female aged between 18-24 years old that included adolescents and youth. The study focused on the universities in each part of Thailand separated as Bangkok, Central, Northeastern, North, and South. This study consists of 15 assistant researchers who are lecturers in each university. The assistant researchers had promoted and facilitated the university students who would like to participate in the study. Moreover, the electronic poster to promote this research had forwarded to university students. Entirely, the number of participants was 500 participants were recruited by convenient sampling.

### The Smart U Health mobile application

The Smart U Health mobile application is a free mobile application developed in the Thai language by the researcher team and runs on the Google Android and iOS operating systems. The participants can download and install via their smartphones. The Smart U Health had shown in the Play Store and App Store at https://play.google.com/store/apps/details?id=th.co. progaming.smartuhealth and https://apps.apple.com/us/app/smart-u-health/id1513492948?l=th&ls=1. It was created between the researcher and the IT team. Information from a previous study and extra information during data collection in the previous study was used to develop Smart U Health. The structure consisted of the first interface to show the project’s name; screening questions consist of 3 questions: the period of smartphone usage, gender, province.

Moreover, this study’s information and informed consent in electronic with the button to click “willing to participate in this study” or “do not want to participate in this study” had included. Then the Smart U Health goes to the next interface: the home interface, which consisted of essential parts to collect the data. It included four parts of the questionnaire:

Part 1: demographic characteristics: age, gender, faculty, year of study, income, chronic diseases

Part 2: the use of smartphones: - the period smartphone usages (years), time spent of smartphone users per day, the purpose of smartphone usages, application usages, location of smartphone usages, charge the battery, eyes rest break

Part 3: the frequency and magnitude of health effects occur while using smartphones after using it during the last three months. There were 15 physical health effects, 12 mental health effects, and 10 social health effects.

Part 4: knowledge, attitudes, and practices regarding the health effects of smartphone usages. The number of questions was 30 in this part.

A self-administered questionnaire had used for data collection throughout the Smart U Health application. The validity of the questionnaire was modified from a previous study [10,18]. Three experts in a related field evaluated the questionnaire, and the validity and reliability were acceptable (IOC=0.90, Cronbach’s alpha=0.75). After thoroughly answering the question, the participants receive the health risk assessment results and the suggestion to reduce health risk levels from smartphone usages.

### Statistical analysis

The licensed SPSS version 22 was used to analyze the data. Frequency, percentage, and mean were used to describe the study population’s general characteristics and study variables. The association between health risk levels and factors had also tested. A bivariate analysis of each vari-able was done first, and then variables with a p-value < 0.05 were included in the multivariate analysis. Statistical analysis was performed using the Statistical Package for the Social Sciences Program (SPSS), version 22. The study protocol was approved by the Ethics Review Committee for Research Involving Human Research Subjects, Health Sciences Group I, Chulalongkorn University (RECCU No.108.1/63).

Risk level = Likelihood x Severity of Consequences

There were ten questions to assess the knowledge regarding the health effects of smartphone usages. A correct answer will receive one score and 0 scores for wrong answers. Attitude and practice scores regarding smartphone usages were classified into three levels using a minimum and maximum interval.

## Results

Of five hundred participants, 328 of them (65.6%) were females, and 172 were males (34.4%). The average age was 20.3±1.5 years old. Two hundred nineteen (43.8%) were studying from faculty related to health. The average income was 7183 baht (218 USD). Most of the participants (91.2%) had no chronic disease.

### The behavior of smartphone usages

The average period of using smartphones among the participants was 7.9±2.0 years, with the average time spent per day was 8.6±3.4 hours. More than half of them (54.8%) used smartphones with an Android operating system, while others used a smartphone with an iOS operating system. The average screen size of their smartphones was 5.2±0.6 inches. Only 18.0 percent of them buy smartphones by themselves. Mainly, the participants used smartphones for making phone calls and applications. Wondering, all of them had the experience of using a smartphone while they were passengers. The top three places that the participants used smartphones were bedroom (98.6%), living room (96.4%), and classroom (95.8%). The top three applications used were reported to be for social networking (99.2%), photo and video recording (98.0%), and working app (76.4%). The participants commonly used their smartphones from 7 pm to 9 pm (89.2%). More than half (58.0%) charge the battery while it dead. Four hundred and eight participants reported that while they were continuously using their smartphones, they sometimes rest their eyes.

### The frequency of health effects from smartphones

The participants reported the top three physical health effects from smartphone usage over three months, including eyes pain (93.8%), shoulder or neck pain (93.6%), and blurred vision (91.0%). The report on the top three mental health effects from smartphone usage included feeling bored (90.4%), lack of concentration (88.4%), and moodiness (87.8%). The top three social health effects from smartphone usage, firstly, the participants had a communication problem with other people (84.0%). Following, reported of lack of concentration while working with others or in the act of doing (81.0%), and strangers’ attraction via social network (81.0%).

**Table I.**
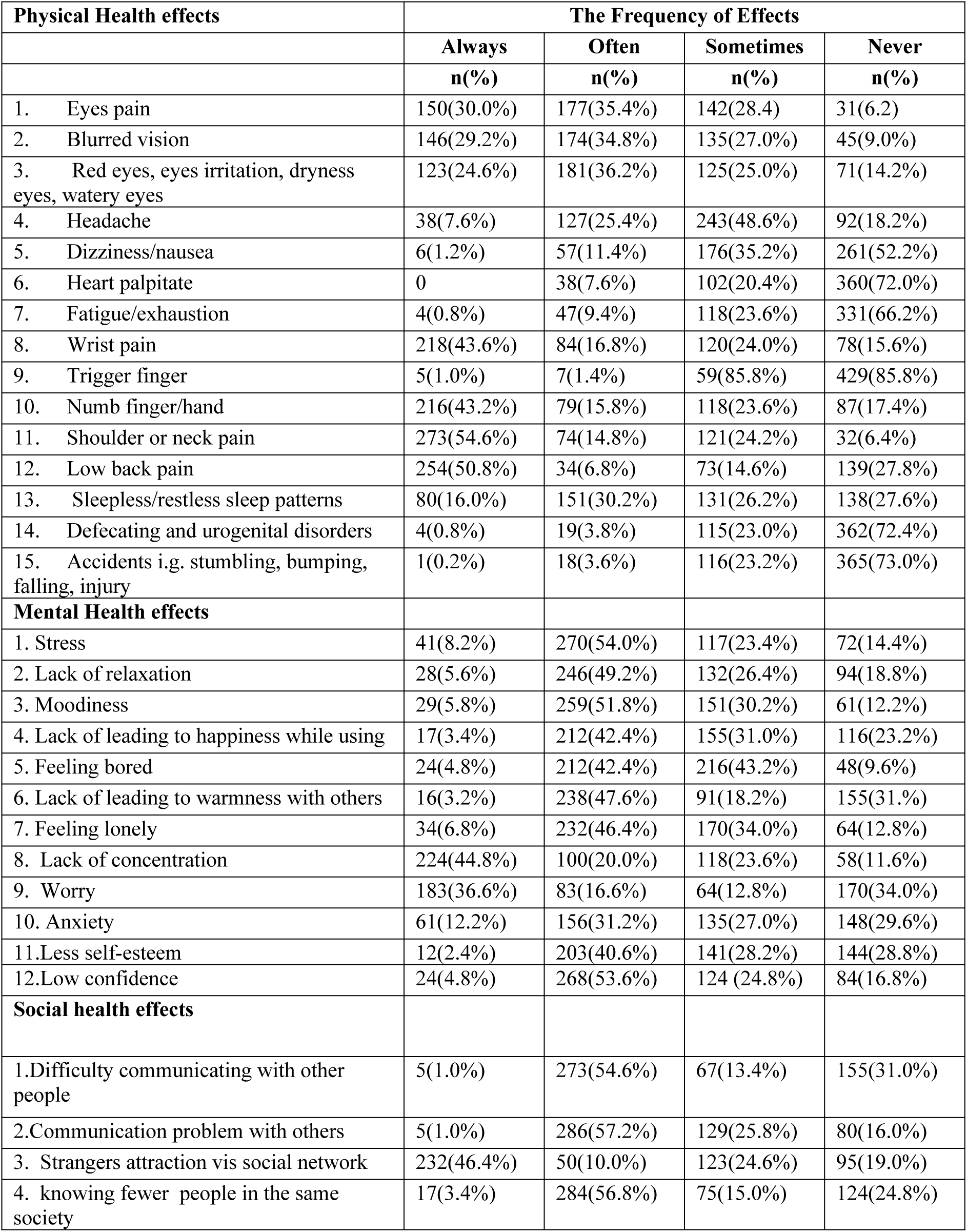
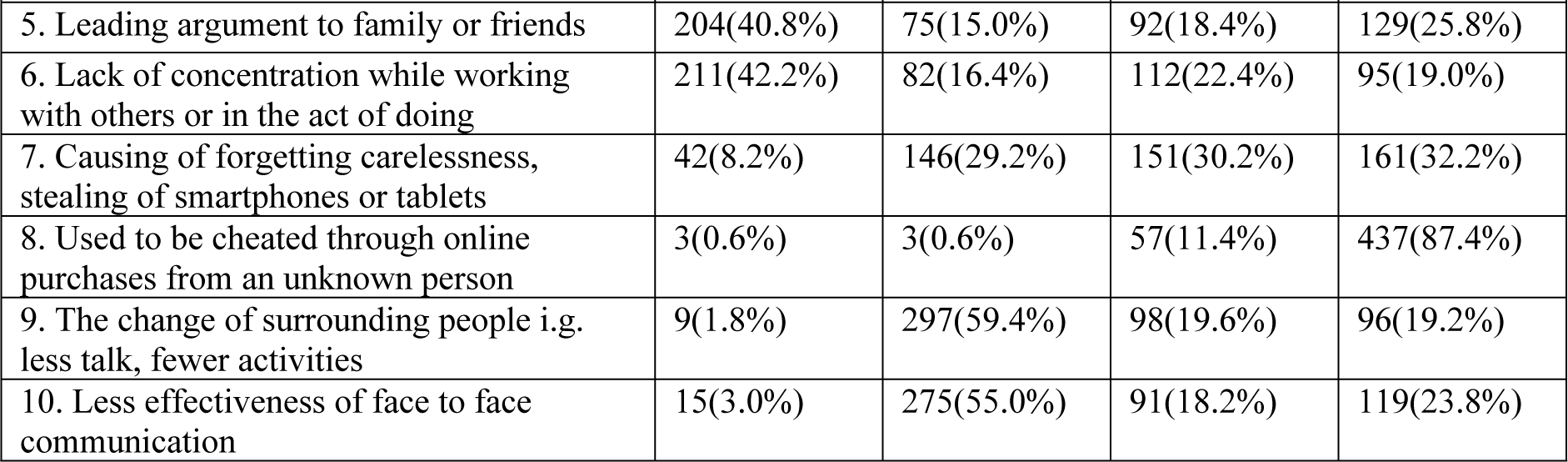
The frequency of health effects from smartphones.

### Association between smartphone usages and health risk levels

A multivariate analysis of factors with health risk levels was run. Participants who had income over 10,000 baht increased physical health risk (OR=3.77; 95%CI=1.16-12.30). Participants who used a smartphone for more than five years had an increase in physical health risk (OR=1.18; 95%CI=1.03-1.35). The participants who mainly used the smartphone for calling had an increased physical health risk (OR=18.85; 95%CI=1.42-250.72). Those who always charge smartphones when they run out of power increased physical health risk (OR=27.06; 95%CI=6.41-114.18). Participants who only sometimes rested their eyes before continuing had an increased physical health risk compared to those who always rest their eyes (OR=2.56; 95%CI=01.20-5.47). Participants who had high knowledge level had an increase in physical health risk level(OR=4.36; 95%CI=1.98-9.60) compared to those who had low or moderate knowledge level while participant who had high practice level had an increase in physical health risk level (OR=5.05; 95%CI=1.12-22.74) compared to those who had low or moderate practice level.

Participants who study in higher years had an increased mental health risk level (OR=2.93; 95%CI=1.21-7.14). Those who study in faculty that in non-health science group had an increased mental health risk level (OR=2.12; 95%CI=1.21-3.74). Participants who had income between 5,001-10,000 baht increased mental health risk (OR=5.20; 95%CI=1.94-13.92). The participants who mainly used the smartphone for calling had an increased mental health risk (OR=17.57; 95%CI=1.12-276.28). Those who always charge smartphones when they run out of power increased mental health risk (OR=24.66; 95%CI=6.92-87.87). Participants who had high knowledge level had an increase in mental health risk level (OR=2.44; 95%CI=1.19-5.00) compared to those who had low or moderate knowledge level while participant who had high practice level had an increase in mental health risk level (OR=6.42; 95%CI=1.75-23.53) compared to those who had low or moderate practice level.

Participants who study in higher years had an increased social health risk level (OR=4.05; 95%CI=1.58-10.34). Those who study in faculty that in non-health science group had an increased social health risk level (OR=2.94; 95%CI=1.49-5.81). Participants who had income between 5,001-10,000 baht increased social health risk (OR=12.31; 95%CI=3.84-39.47). Participants who used smartphones in the classroom had an increased social health risk level (OR=8.75; 95%CI=1.95-39.35). Participants who used work application had an increased social health risk level (OR=2.07; 95%CI=1.00-4.27). Those who always charge smartphones when they run out of power increased social health risk (OR=93.98; 95%CI=16.59-532.37). Participants who never rested their eyes before continuing had an increased social health risk compared to those who always rest their eyes (OR=16.46; 95%CI=2.26-119.93). Participants who had high knowledge level had an increase in social health risk level(OR=2.58; 95%CI=1.05-6.34) compared to those who had low or moderate knowledge level while participant who had high practice level had an increase in social health risk level (OR=13.71; 95%CI=1.49-125.72) compared to those who had low or moderate practice level.

**Table II.**
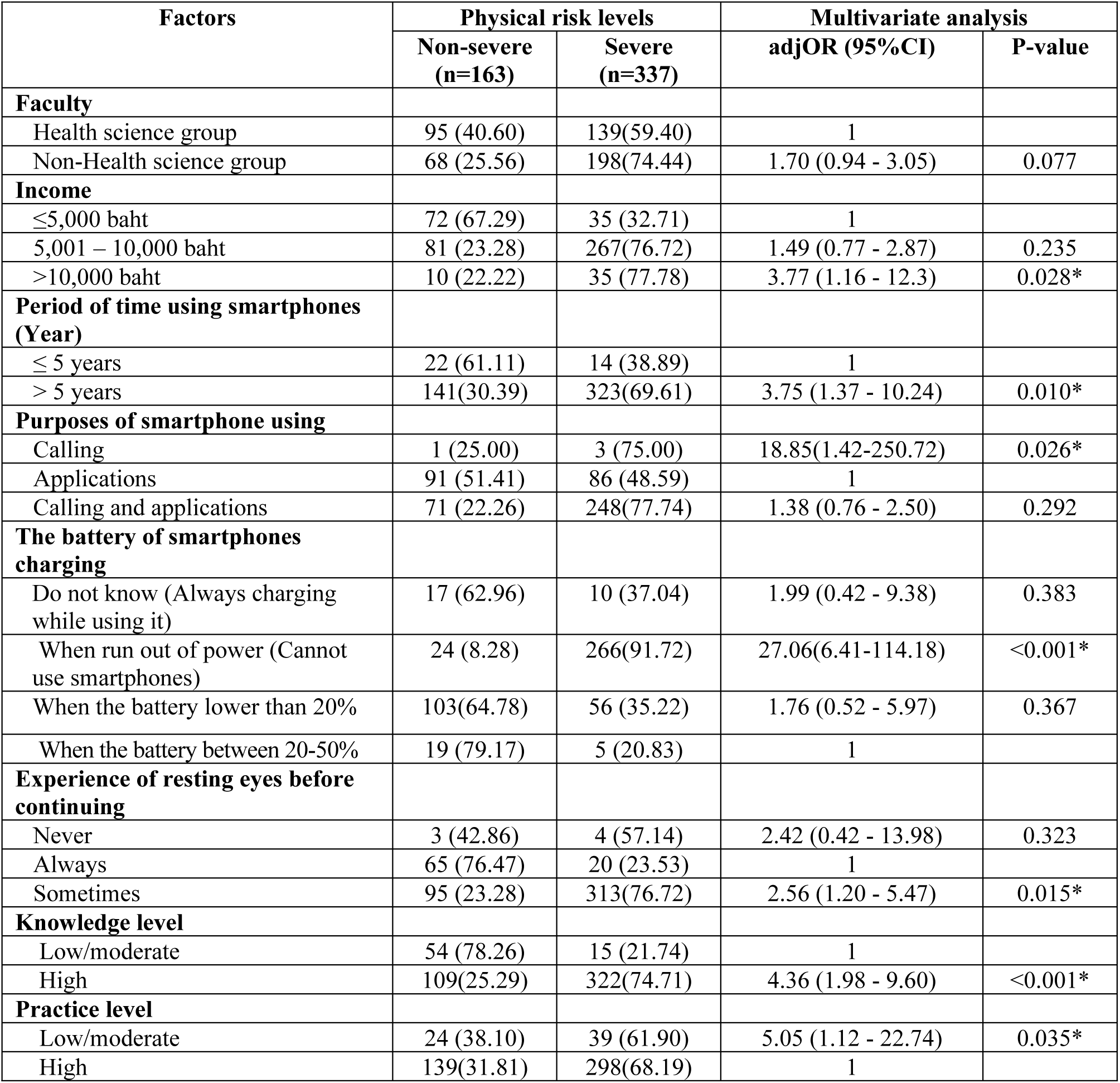
Multivariate analysis of factors with physical risk level among university students in Thailand (n = 500)

**Table III.** Multivariate analysis of factors with mental risk level among university students in Thailand (n = 500)

| Factors | Mental risk levels |  | Multivariate analysis |  |
| --- | --- | --- | --- | --- |
|  | Non-severe<br>(n=163) | Severe<br>(n=337) | adjOR (95%CI) | P-value |
| **Study level** |  |  |  |  |
| Year 1 -4 | 140 (33.18) | 282 (66.82) | 1 |  |
| Year 5 -6 | 23 (29.49) | 55 (70.51) | 2.93 (1.21 - 7.14) | 0.018* |
| **Faculty** |  |  |  |  |
| Health science group | 99 (42.31) | 135 (57.69) | 1 |  |
| Non-Health science group | 64 (24.06) | 202 (75.94) | 2.12 (1.21 - 3.74) | 0.009* |
| **Income** |  |  |  |  |
| ≤5,000 baht | 23 (63.89) | 13 (36.11) | 1 |  |
| 5,001 – 10,000 baht | 140 (30.17) | 324 (69.83) | 5.20 (1.94 - 13.92) | 0.001* |
| **Purposes of smartphone using** |  |  |  |  |
| Calling | 1 (25.00) | 3 (75.00) | 17.57(1.12-276.28) | 0.041* |
| Applications | 94 (53.11) | 83 (46.89) | 1 |  |
| Calling and applications | 68 (21.32) | 251 (78.68) | 1.64 (0.92 - 2.92) | 0.093 |
| **The battery of smartphones charging** |  |  |  |  |
| Do not know (Always charging while using it) | 17 (62.96) | 10 (37.04) | 1.90 (0.47 - 7.69) | 0.371 |
| When run out of power (Cannot use smartphones) | 27 (9.31) | 263 (90.69) | 24.66(6.92-87.87) | <0.001* |
| When the battery lower than 20% | 101 (63.52) | 58 (36.48) | 1.79 (0.59 - 5.47) | 0.306 |
| When the battery between 20-50% | 18 (75.00) | 6 (25.00) | 1 |  |
| **Knowledge level** |  |  |  |  |
| Low/moderate | 50 (72.46) | 19 (27.54) | 1 |  |
| High | 113 (26.22) | 318 (73.78) | 2.44 (1.19 – 5.00) | 0.015* |
| **Practice level** |  |  |  |  |
| Low/moderate | 23 (36.51) | 40 (63.49) | 6.42 (1.75-23.53) | 0.005* |
| High | 140 (32.04) | 297 (67.96) | 1 |  |

**Table IV.** Multivariate analysis of factors with social risk level among university students in Thailand (n = 500)

| Factors | Social risk levels |  | Multivariate analysis |  |
| --- | --- | --- | --- | --- |
|  | Non-severe (n=190) | Severe (n=310) | adjOR (95%CI) | P-value |
| **Study level** |  |  |  |  |
| Year 1 -4 | 165(39.10) | 257(60.90) | 1 |  |
| Year 5 -6 | 25 (32.05) | 53 (67.95) | 4.05 (1.58 - 10.34) | 0.004* |
| **Faculty** |  |  |  |  |
| Health science group | 114(48.72) | 120(51.28) | 1 |  |
| Non-Health science group | 76 (28.57) | 190(71.43) | 2.94 (1.49 - 5.81) | 0.002* |
| **Income** |  |  |  |  |
| ≤5,000 baht | 27 (75.00) | 9 (25.00) | 1 |  |
| 5,001 – 10,000 baht | 163(35.13) | 301(64.87) | 12.31 (3.84 - 39.47) | <0.001* |
| **Purposes of smartphone using** |  |  |  |  |
| Calling | 71 (78.89) | 19 (21.11) | 9.31 (0.42-207.33) | 0.159 |
| Applications | 114(28.22) | 290(71.78) | 1 |  |
| Calling and applications | 5 (83.33) | 1 (16.67) | 1.84 (0.96 - 3.53) | 0.068 |
| **Using smartphones in classroom** |  |  |  |  |
| No | 16 (76.19) | 5 (23.81) | 1 |  |
| Yes | 174(36.33) | 305(63.67) | 8.75 (1.95 - 39.35) | 0.005* |
| **Using work applications** |  |  |  |  |
| No | 69 (58.47) | 49 (41.53) | 1 |  |
| Yes | 121(31.68) | 261(68.32) | 2.07 (1.00 - 4.27) | 0.05* |
| **The battery of smartphones charging** |  |  |  |  |
| Do not know (Always charging while using it) | 22 (81.48) | 5 (18.52) | 1.54 (0.21-11.35) | 0.673 |
| When run out of power (Cannot use smartphones) | 28 (9.66) | 262(90.34) | 93.98(16.59-532.37) | <0.001* |
| When the battery lower than 20% | 119(74.84) | 40 (25.16) | 3.16 (0.66-15.08) | 0.148 |
| When the battery between 20-50% | 21 (87.50) | 3 (12.50) | 1 |  |
| **Experience of resting eyes before continuing** |  |  |  |  |
| Never | 3 (42.86) | 4 (57.14) | 16.46 (2.26-119.93) | 0.006* |
| Always | 70 (82.35) | 15 (17.65) | 1 |  |
| Sometimes | 117(28.68) | 291(71.32) | 1.61 (0.67 - 3.84) | 0.283 |
| **Knowledge level** |  |  |  |  |
| Low/moderate | 56 (81.16) | 13 (18.84) | 1 |  |
| High | 134(31.09) | 297(68.91) | 2.58 (1.05 - 6.34) | 0.038* |
| **Practice level** |  |  |  |  |
| Low/moderate | 28 (44.44) | 35 (55.56) | 13.71(1.49-125.72) | 0.021* |
| High | 162(37.07) | 275(62.93) | 1 |  |

## Discussions

This current study is the first study investigating the factors associated with health risk level from smartphone usage among university students in Thailand via mobile application. The average smartphone usage in this current study among university student users was higher than the average among older adults in Thailand, which reported the average (±SD) time spent on smartphones was 2.7 (±1.5) hours/day [18]. However, the Thai students used their smartphones for almost the same time as Malay students (8.87 ± 5.16) [22]. Social networking applications were popular among the participants in this study, similarly to a previous study among students in Japan who reported they mostly used Facebook, Twitter, LINE [23]. The university students in this current study had experience using smartphones in the classroom with support from another study in Korea. The average used was 21.06 min on average (out of 75-min effective class duration) among college students in Korea [14].

The reported physical, mental, and social health effects in this study consisted of many previous studies. A previous case-control study found that smartphone usage is a risk factor for pediatric dry eye disease among children and adolescents [24]. Moreover, other studies also reported that their participants had headaches or dizziness from using smartphones [25]. Some of the currents study participants reported that they had to experience respiratory function problems, which, similarly to a previous study, showed an increase in the adverse effects on respiratory functions when using smartphones for more than 4 hours [26]. More support from other studies that the use of smartphones is a risk factor for the musculoskeletal problem [27], hand pain [14], and neck problems [13]. Moreover, a previous study supported that smartphone usage affected sleep quality [28]. The report of poor mental health from smartphones in this current study consisted of a previous study [29]. Increasing anxiety, depression, and stress were reported from smartphone users [30]. Additional studies supported the current study that the use of smartphones increased emotional problems, lower self-esteem [31], social anxiety, and loneliness [32]. Social health effects from the use of smartphones in this study, similarly to the previous study among university students in Thailand, found that over the use of smartphones had lower scores on the social-psychological well-being than those who did not (P < 0.001) [33]. Another study showed that overused smartphones among students had lower communication quality with their parents [31]. Moreover, adolescents reported cyber victimization and cyberbullying by using smartphones [34].

The factors associated with a health risk from smartphone usage in the current study consistent with other studies. Income was a factor associated with a health risk from smartphone usage in the current study support by the results from another study also showed an association between smartphone usage and family income [35]. The current study similarly to the study in Korea, the results found a significant difference between the duration of smartphone usage and the craniovertebral angle, scapular index, and peak expiratory flow related to show negatively affect both posture and respiratory function [26]. Another study in Japan similar to the current study. Many applications were popular, but it is coming with time spent in the real-life decreasing. Online gaming popular among males, while females use the internet for communications online. Those activities affected mental health. Therefore, smartphone users should be aware of the seriousness of internet addictions [23]. The result on battery charging in the current study can assume that it is a similar way to predict smartphones’ overuse because some smartphone users will charge the battery of the smartphones only when it cannot use. A problematic smartphone mainly uses analysis from a longer daily smartphone screen time and evening chronotype personality from a previous study. The consistency between the current study and a previous one showed a strong association between too much screen time and the problematic smartphone use level (Pavle, 2019). A previous study found consistent results that age at first using a smartphone, duration on the smartphone per day (≥ 9 hours), and depression carried a higher risk of developing problematic smartphone use. However, the results from the current study inconsistent because a previous study showed that the field of study (science vs. arts-based) did not contribute to an increased risk of developing problematic smartphone use [36]. Using smartphones in the classroom was an associated factor with health risk level similar to the study in 2019, which found that the students used their phones for more than 20 min within a class duration. Daily and in-class use of the phone negatively affected student grades [37]. It may assume that using a smartphone in the classroom affect the user’s concertation on the study. Another study on dry eye disease showed that more prolonged daily smartphone use might be a risk factor. The user in the older year of study and older-aged years show a higher prevalence of dry eye disease. Besides, dry eye disease prevalence was lower in older grade children from rural groups compared to younger grade children in urban groups [24]. Another study showed many factors associated with social anxiety or loneliness, such as receiving fewer incoming calls and using healthy applications more frequently. Lonely individuals tend to use the system, beautify, browser, and social media (RenRen) apps more frequently, while Individuals with higher levels of social anxiety also receive less SMSs and use camera apps less frequently [32].

## Conclusion

The smartphone users had many health effects on smartphone usage. The Smart U Health mobile application might be the appropriate tool to assess health risk levels among smartphone users. In the future, may consider intervention program to give awareness to smartphone users.

## Data Availability

All relevant data are within the manuscript and its Supporting Information files.

## Acknowledgements

The authors are thankful to the Ratchadapiseksomphot Fund, Chulalongkorn University for the Postdoctoral Fellowship, funded by Chulalongkorn University (CU_GI_63_08_53_01), the grant for International Research Integration: Chula Research Scholar (GCURS_59_06_79_01) Ratchadaphiseksomphot Endowment Fund and the Office of International Affairs Scholarship for Short-term Research, Chulalongkorn University.

